# Patterns of Gabapentin Use in Patients With Cervical Spondylotic Myelopathy

**DOI:** 10.64898/2026.07.27.26358977

**Authors:** Benjamin C. Warner, Faraz Arkam, Salim Yakdan, Ahmad Hammo, Wilson Z. Ray, Adam Wilcox, Randi Foraker, Chenyang Lu, Jacob K. Greenberg

## Abstract

Cervical spondylotic myelopathy (CSM) is the most common cause of nontraumatic spinal cord dysfunction in adults and is an increasingly important source of disability as populations age. Gabapentin is widely prescribed for neuropathic pain and may therefore be used for symptoms related to known or undiagnosed CSM. However, there is sparse evidence related specifically to gabapentin’s use for CSM-related pain. We investigate gabapentin use and trends over time in patients with CSM compared to matched controls. We observed that gabapentin prescriptions were higher in CSM patients compared to controls across two multi-hospital datasets. These results highlight the need for further research into pharmacologic treatment for chronic pain in CSM.

## Introduction

Cervical spondylotic myelopathy (CSM) is the most common cause of nontraumatic spinal cord dysfunction in adults and is an increasingly important source of disability as populations age.^1^ Pain is among the most prevalent symptoms of CSM, reported by roughly 80% of patients at presentation.^2^ Although substantial attention has been focused on surgical management and postoperative outcomes, far less evidence guides pharmacologic treatment of these patients.^3^

Gabapentin is widely prescribed for neuropathic pain and may therefore be used for symptoms related to known or undiagnosed CSM.^4^ However, there is sparse evidence related specifically to gabapentin’s use for CSM-related pain.^3^ Evidence supporting gabapentin for nonspecific chronic spinal pain is limited, with most studies showing no benefit or only small effect sizes.^5,6^ Moreover, adverse effects such as dizziness, somnolence, ataxia, and gait disturbance are common and may complicate neurologic assessment of CSM severity and progression.^4^ We investigate gabapentin use and trends over time in patients with CSM compared to matched controls.

## Methods

We performed a retrospective study using structured electronic health record data from Washington University/BJC HealthCare and administrative claims from the Merative MarketScan database. In the BJC cohort, 13,200 patients with CSM were matched to 419,189 controls; in the Merative cohort, 30,772 CSM cases were matched to 1,445,438 controls. Cases and controls were matched at an approximate ratio of 35:1 to emulate current prevalence estimates of CSM in the general population.^7^ CSM was matched using ICD-10 and corresponding SNOMED codes mapped through the Athena database and FEMR.^7,8^ We identified first prescriptions analgesic agents from 1 year before to 1 year after the index date, defined as the confirming CSM-related diagnosis for cases and the matched date for controls.

## Results

Among patients who received any temporally-relevant prescription in the BJC cohort, as seen in Figure 1, gabapentin prevalence as a first prescription among CSM patients rose from 21.8% in 2011 to 33.5% in 2023, whereas rates in matched controls declined marginally from 15.7% to 14.6% concurrently. In the Merative cohort from 2012 to 2021, gabapentin rose substantially among CSM cases (16.0% to 25.0%) and increased from 9.1% to 16.5% in the controls. Among all patients regardless of prescription status, as indicated in Figure 2, we observed a 0.82%/year (95% CI: 0.38-1.26, □ = *1.23* * *10*^−*3*^) difference between CSM patients and controls (1.545%/year, □^*2*^ = *0.860* vs. 0.727%/year, □^*2*^ = *0.858*, respectively) in gabapentin prevalence in the BJC dataset; and observed a 0.89%/year (95% CI: 0.32-1.47, □ = *5.43* * *10*^−*3*^) difference between CSM patients and controls (1.394%/year, □ ^2^ = *0.815* vs. 0.501%/year, □^*2*^ = *0.686*, respectively) in gabapentin prevalence in the Merative dataset.

**Fig. 1.**
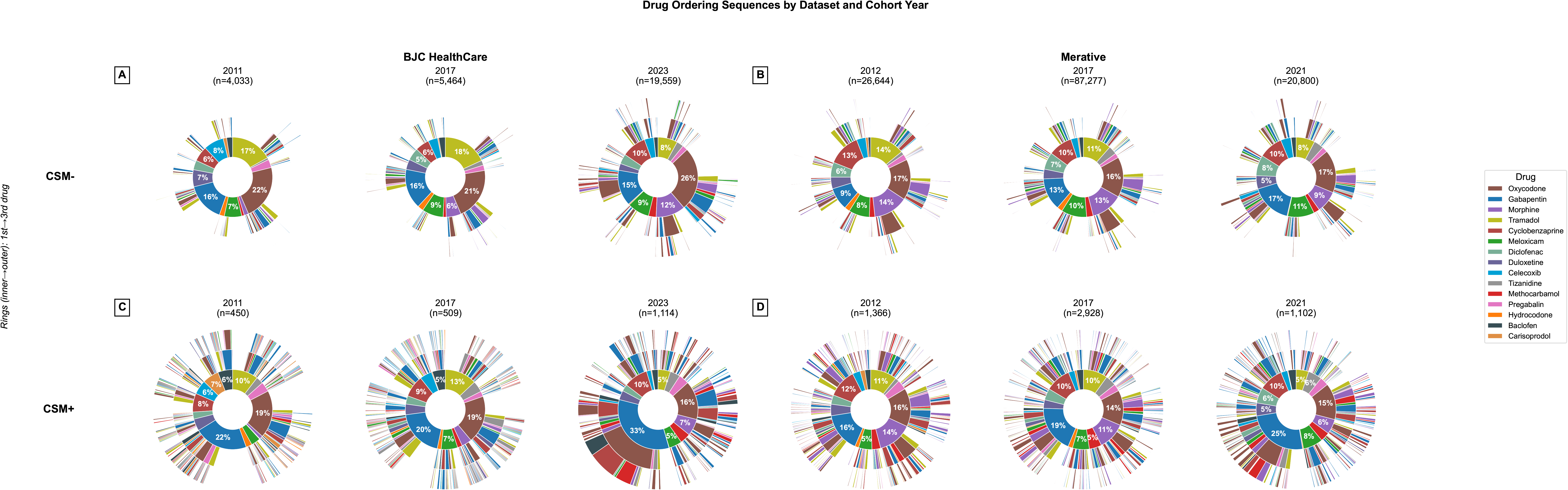

**Fig. 2.**
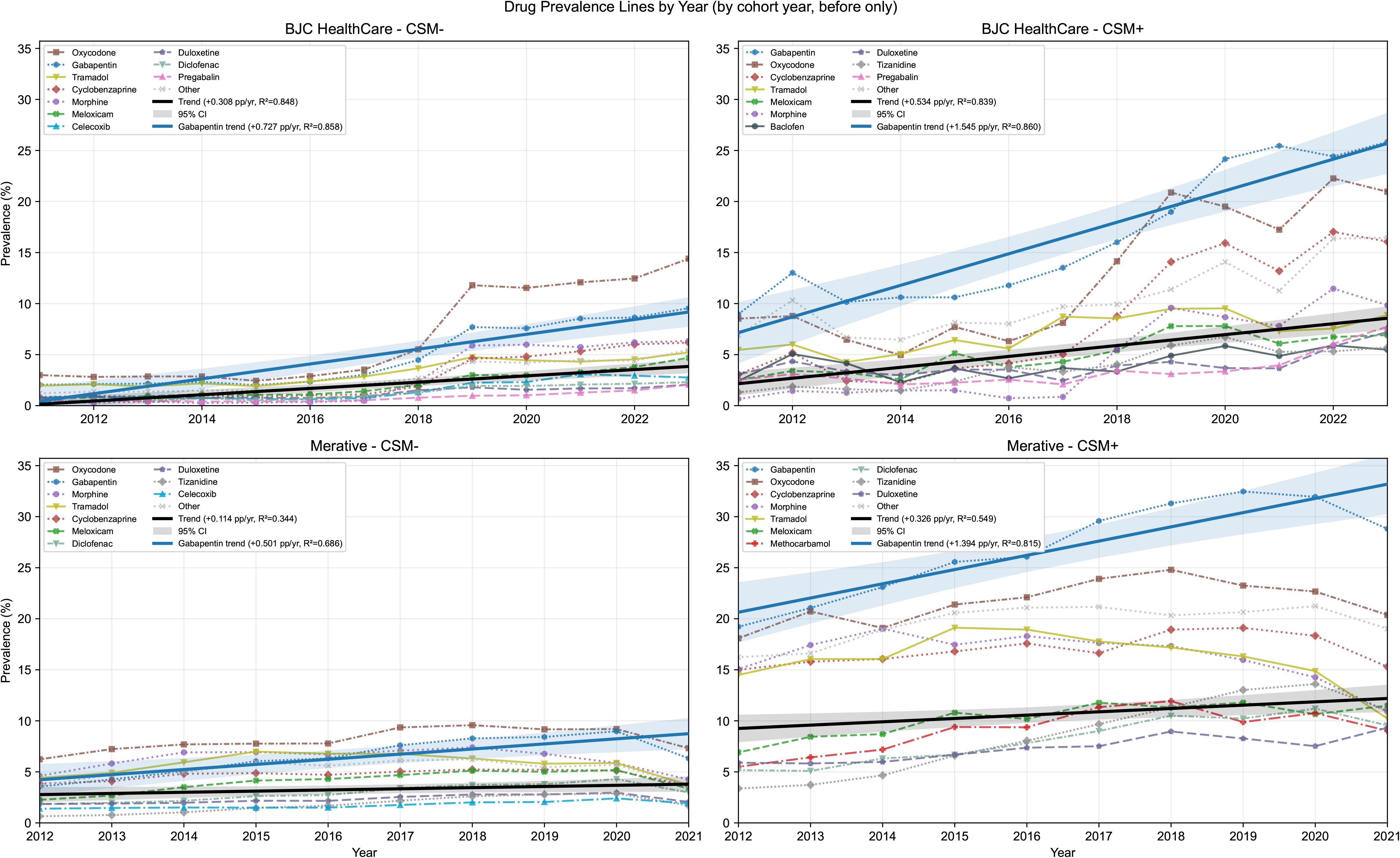

Aggregated over all years, gabapentin was the most common analgesic among CSM cases. In BJC, the 3 most common pre-index analgesics among CSM cases were gabapentin (16.9%), oxycodone (13.3%), and cyclobenzaprine (9.0%), compared with oxycodone (7.2%), gabapentin (5.1%), and tramadol (3.4%) among controls. In Merative, the 3 most common pre-index analgesics among CSM cases were gabapentin (27.1%), oxycodone (21.9%), and cyclobenzaprine (17.1%), compared with oxycodone (8.3%), gabapentin (6.8%), and morphine (6.5%) among controls.

We also observe that among prescriptions, gabapentin prescriptions were more proximate to the index. In the BJC cohort, the mean interval from first gabapentin prescription to the index was 138 days (95% CI: 134-142) among CSM cases compared with 225 days (95% CI: 224-226) among controls. In the Merative cohort, the corresponding intervals were 171 days (95% CI: 168-173) for CSM cases and 209 days (95% CI: 209-210) for controls.

## Discussion

We observed that gabapentin prescriptions were higher in CSM patients compared to controls across two multi-hospital datasets. One possible explanation is that gabapentin is being prescribed for pain or numbness/paresthesias arising from unrecognized cervical cord compression, which is possible given that CSM is frequently identified late in its clinical course.^9,10^ Alternatively, CSM may be known but uncoded, with clinicians treating CSM-related pain unknowingly, or speculatively so with limited supporting evidence.

While these explanations have different implications, the persistent use post-CSM diagnosis may have significant consequences. Somnolence, cognitive changes, and gait disturbance may confound CSM assessments in a population already impacted by balance and gait impairment.^4,9^ With both the risk and benefit of gabapentin being poorly defined in this population, appropriate guidance for patients remains unclear. Given that pain is a principal driver of both patient presentation and management decisions in CSM, prospective studies are needed to evaluate the role, effectiveness, and safety of gabapentin in this population.^2,3^

## Data Availability

The BJC HealthCare data is not publicly available, but may be made available upon request with an appropriate data use agreement. The Merative MarketScan Research Databases are available through a commercial license with Merative.

## Acknowledgements

The authors would like to thank Shinji Naka for his efforts in preparing the BJC HealthCare data, as well as Charles Alba for reviewing the code utilized in this study. The authors would also like to thank all of the patients involved in furthering the understanding of cervical spondylotic myelopathy.

## Data Sharing Statement

The BJC HealthCare data is not publicly available, but may be made available upon request with an appropriate data use agreement. The Merative® MarketScan™ Research Databases are available through a commercial license with Merative.

## References

1. Yakdan S, Joseph K, Poulin N, et al. Disparities in quality of life and health literacy among patients with degenerative cervical myelopathy: the influence of racial and ethnic factors in the All of Us Research Program. Journal of Neurosurgery: Spine. 2026;44(6):920–929. doi:10.3171/2025.11.SPINE25662.

2. Schneider MM, Badhiwala JH, Alvi MA, et al. Prevalence of neck pain in patients with degenerative cervical myelopathy and short-term response after operative treatment: a cohort study of 664 patients from 26 global sites. Glob Spine J. 2024;14(3):830–838. doi:10.1177/21925682221124098.

3. Levett JJ, Georgiopoulos M, Martel S, et al. Pharmacological treatment of degenerative cervical myelopathy: a critical review of current evidence. Neurospine. 2024;21(2):375–400.

4. CADTH. Gabapentin for Adults with Neuropathic Pain: A Review of the Clinical Efficacy and Safety. Canadian Agency for Drugs and Technologies in Health; 2015.

5. Atkinson JH, Slater MA, Capparelli EV, et al. A randomized controlled trial of gabapentin for chronic low back pain with and without a radiating component. Pain. 2016;157(7):1499–1507.

6. Shanthanna H, Gilron I, Rajarathinam M, et al. Benefits and safety of gabapentinoids in chronic low back pain: a systematic review and meta-analysis of randomized controlled trials. PLoS Med. 2017;14(8):e1002369.

7. Yakdan S, Warner B, Ghogawala Z, et al. Clinically-guided models or foundation models? Predicting cervical spondylotic myelopathy from electronic health records. npj Digit Med. 2026;9:153.

8. Steinberg E, Fries JA, Xu Y, Shah N. MOTOR: A Time-to-Event Foundation Model For Structured Medical Records. In: The Twelfth International Conference on Learning Representations. 2024. https://openreview.net/forum?id=NialiwI2V6

9. The Lancet Neurology. A focus on patient outcomes in cervical myelopathy. Lancet Neurol. 2019;18(7):615.

10. Asuzu DT, Yun JJ, Alvi MA, et al. Association of ≥12 months of delayed surgical treatment for cervical myelopathy with worsened postoperative outcomes: a multicenter analysis of the Quality Outcomes Database. J Neurosurg Spine. 2022;36(4):568–574. doi:10.3171/2021.7.SPINE21590

