## Supplementary Materials for "Patterns of Gabapentin Use in Patients With Cervical Spondylotic Myelopathy"

**eMethods**. We utilize EHRs collected from the Merative MarketScan Commercial Database and

institutionally derived EHRs drawn from Washington University/BJC HealthCare in St. Louis,

Missouri. Patient data from the Washington University/BJC HealthCare system was obtained under a retrospective waiver of consent (IRB #202306053). Patients were collected in a matched control-cohort group of cervical spondylotic myelopathy patients, the latter of which were selected if they had at least two diagnoses of ICD-10 M47.11-M47.13 or M50.0x or their ICD-9 equivalents. Control patients were matched to have a corresponding primary care visit within 30 days of the matched cohort patient’s diagnosis and had an age within 10 years of the cohort patient.

We utilize a mixture of Polars 1.37.1 and Pandas 2.3.3 in our analysis with Python 3.12.12. Selected data was queried from Azure Databricks into sharded Parquet files for all medications, diagnoses, and surgical procedures, and subsequently analyzed through aggregation. Claude Code by Anthropic was used to generate the majority of the code utilized in the analysis; and was subsequently verified by the author B.C.W. The code utilized here is available at <https://github.com/bcwarner/csm-painkillers>.
